# Uterine Fibroids Increase Stroke Risk: a Mendelian Randomization Study

**DOI:** 10.1101/2025.03.20.25324360

**Authors:** Ming Zheng

**Affiliations:** Beijing Institute of Basic Medical Sciences, 27 Taiping Road, Beijing 100850, China; Academy of Military Medical Sciences, 27 Taiping Road, Beijing 100850, China

**Keywords:** Uterine fibroids, Ischemic stroke, Mendelian randomization, Genome-wide association study, Single nucleotide polymorphism

## Abstract

Uterine fibroid, also called leiomyomas, is the most common tumor in women, affecting more than 70% of women over the age of 50. As a generally benign disease, the uterine fibroid can potentially contribute to female stroke through different pathological conditions. The relationship between uterine fibroids and stroke was occasionally reported in sporadic case reports. However, fibroids-related stroke still remains elusive at the population level. This study conducted Mendelian randomization (MR) analysis to investigate the risk of stroke (outcome) following uterine fibroid (exposure). Furthermore, through interrogating the largest clinically representative cohort of 521,612 participants, this study had sufficient power to detect the causality of uterine fibroid on different stroke subtypes, thus discovering the small vessel ischemic stroke (IS) as the most significant stroke subtype associated with uterine fibroids. In conclusion, this study explicitly indicates that uterine fibroids can causally increase the risk of stroke, rather than just a benign disease. This finding strongly suggests the importance of encompassing female populations in stroke studies and the investigation of fibroid-related stroke as a novel stroke subtype from animal models through clinical trials.

**Highlights:**

- Uterine fibroids can causally increase the risk of stroke, rather than just a benign disease.
- Small vessel ischemic stroke is the most significant stroke subtype associated with uterine fibroids.

## Introduction

Uterine fibroid, also called leiomyomas, is the most common tumor in women, affecting more than 70% of women over the age of 50.[1] Uterine fibroids are usually benign and often present without noticeable symptoms.[2] Among women with fibroids, approximately 15 to 30% of them will develop mild but bothersome symptoms, including abdominal pain, excessive menstrual bleeding, and anemia.[3] In summary, the uterine fibroid is a benign gynecological disease that may reduce the health-related quality of life.[4]

As a generally benign disease, the uterine fibroid can potentially contribute to female stroke by different pathological conditions: **(1)** the embolization caused by the thrombosis of adjacent veins compressed by enlarged fibroids;[5, 6] **(2)** the thrombocytosis, hypercoagulable state, and anemic hypoxia caused by the iron deficiency anemia due to bleeding from fibroids;[7, 8] **(3)** the recurrent cerebral infarction that can be treated by the surgical removal of fibroids.[9] The relationship between uterine fibroids and stroke was occasionally reported in sporadic case reports.[5, 6, 7, 8, 9] However, fibroids-related stroke still remains elusive at the population level.

## Methods

To investigate the causal inference from uterine fibroid to stroke, this study conducted Mendelian randomization (MR) analysis to investigate the genetically determined fibroids in a stroke cohort of 521,612 participants.[10] Using data from a genome-wide association study (GWAS) of a uterine fibroid cohort of 123,579 participants,[11] genetic instruments of single nucleotide polymorphisms (SNPs) for fibroids were selected using a *p*-value <5×10^−7^ and an independent inheritance with a minimal level of linkage disequilibrium (LD) of r^2^ <0.001. Instrumental strength was quantified using the *F*-statistic. A total of 80 SNPs were selected with an average *F*-statistic of 56.5 (range: 25.2-307.9; **Figure. 1A**), with an *F*-statistic >10 being considered sufficiently informative.[12] This study applied four MR methods, including simple mode,[13] weighted median[14], weighted mode[13], and inverse-variance weighting (IVW)[12]. Causal estimate was calculated by log-odds ratio. MR pleiotropy residual sum and outlier (MR-PRESSO) test was conducted to correct the horizontal pleiotropy when the MR-PRESSO global test *p*-value was <0.05.[15] Multiple testing correction was performed using the false discovery rate (FDR) method.[16]

**Figure 1.**
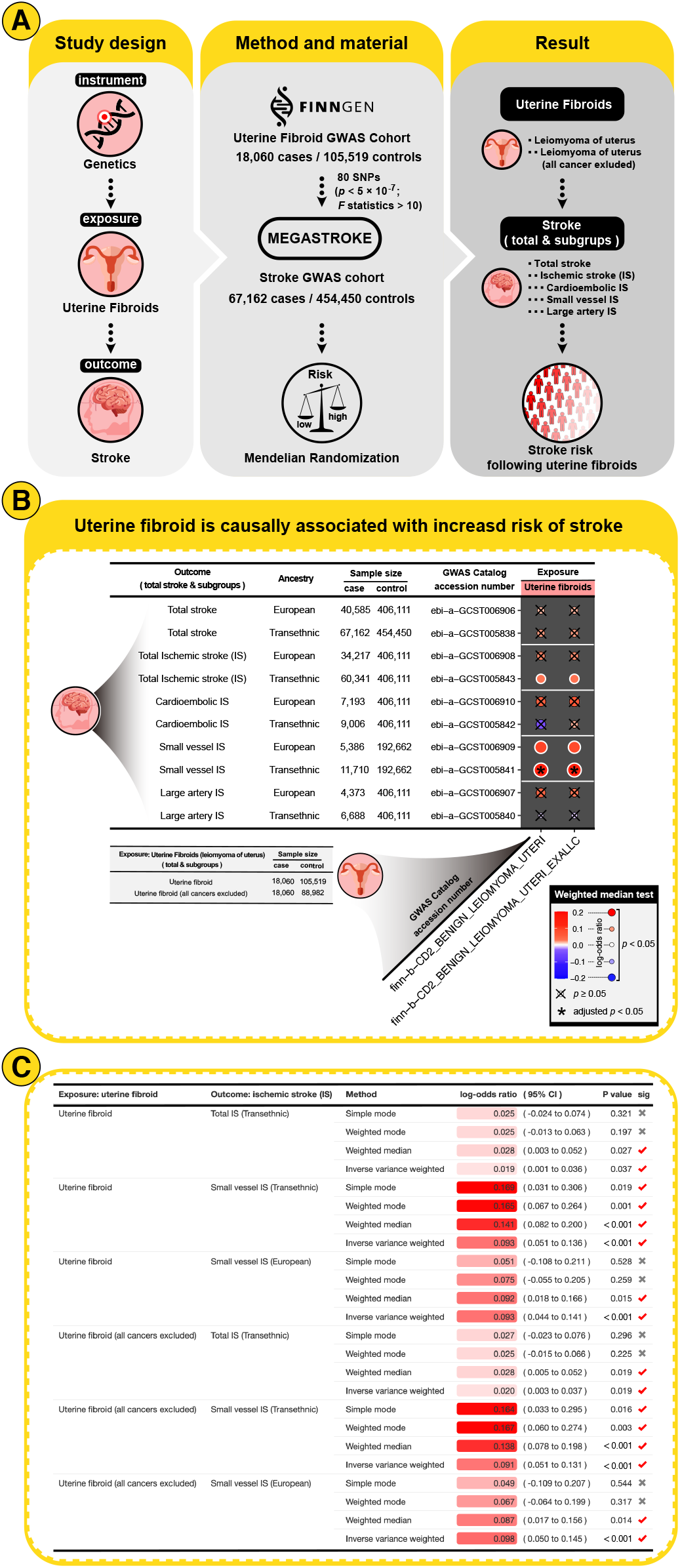
Mendelian randomization study of uterine fibroids and stroke. **(A)** The graphical illustration of the mendelian randomization (MR) study. The MR analysis was conducted using the genetic instrument to estimate the causal role of uterine fibroids exposure in stroke outcomes. This MR analysis tested the 80 genetic variants of uterine fibroids against stroke in genome-wide association study (GWAS) cohorts to evaluate the stroke risk following uterine fibroids. **(B)** MR results show the causal effect of uterine fibroids on the risk of stroke. MR analysis estimated causal effects were calculated using the weighted median test, with horizontal pleiotropy corrected when MR pleiotropy residual sum and outlier (MR-PRESSO) Global test *p* <0.05. The causal estimates were shown by heatmap, with the dot color and size representing the log-odds ratio. A positive value of log-odds ratio indicates that uterine fibroid is associated with an increased risk of stroke. The “×” symbol represents false discovery rate (FDR)-adjusted *p* ≥ 0.05; and the “*” symbol represents adjusted *p* <0.05. **(C)** Causal estimates of genetically determined uterine fibroids on the risk of ischemic stroke (IS). Causal estimates were analyzed by different MR methods, including simple mode, weighted median, weighted mode, and inverse-variance weighting (IVW). The MR causal effect was shown by the color gradient. The significance of *p* <0.05 were indicated by “√” symbol.

## Results

This MR study investigated the risk of stroke (outcome) following uterine fibroid (exposure). As shown in **Figure. 1B**, uterine fibroids were significantly associated with increased risks of IS, including total IS and small vessel IS (FDR-adjusted *P*_weighted median_ <0.05). The most significant causality was found between uterine fibroids and small vessel IS (causal estimate =0.14; *P*_MR-PRESSO_ <0.001).

Furthermore, the robustness of the MR results was investigated using different MR methods: simple mode,[13] weighted median[14], weighted mode[13], and inverse-variance weighting (IVW)[12]. The causal effect of uterine fibroid on small vessel IS was significant across different MR tests (**Figure. 1C**). Additionally, the causalities of uterine fibroid on total IS were significant in the IVW and weighted median tests but not in the simple mode and weighted mode tests. Nevertheless, there was a universal trend toward increased IS risks following uterine fibroid exposure **(Figure. 1C)**.

## Discussion

To the best of our knowledge, this is the first MR study demonstrating the causal inference from uterine fibroid to increased stroke risk. Through interrogating the largest clinically representative cohort of stroke, this study had sufficient power to detect the causality of uterine fibroid on different stroke subtypes, thus discovering the small vessel IS as the most significant stroke subtype associated with uterine fibroids. Thus, different subtypes of stroke should be considered in future research.

Of note, the true prevalence of uterine fibroids may not be accurately determined, because the majority (~70%) of women with uterine fibroids had no symptoms.[3] Therefore, the studies of uterine fibroids are potentially tainted by potential confounders. Hopefully, this study analyzed genetically determined uterine fibroids by genetic variants, which are less susceptible to confounding factors or reverse causation.[17]

Given the causality of uterine fibroids on stroke, it is reasonable to assume that the treatment of uterine fibroids might reduce the risk and severity of stroke. In a case report, the recurrence of ischemic stroke can be controlled by the surgical removal of uterine fibroids.[9] Fibroid-related stroke could be attributed to the compression of adjacent veins and fibroid-related thrombocytosis and anemia.[5, 6, 7, 8] Future research is warranted to explore the mechanisms of fibroid-related stroke. More insight into the underlying mechanisms is important for the development of female-specific strategies for the treatment and prevention of stroke.

## Conclusion

This study explicitly indicates that uterine fibroids can causally increase the risk of stroke, rather than just a benign disease. This finding strongly suggests the importance of encompassing female populations in stroke studies and the investigation of fibroid-related stroke as a novel stroke subtype from animal models through clinical trials.

## Data Availability

All data produced in the present study are available upon reasonable request to the authors.

## Nonstandard Abbreviations and Acronyms

IS: ischemic stroke
MR: Mendelian randomization
MR-PRESSO: Mendelian randomization pleiotropy residual sum and outlier
GWAS: genome-wide association study
SNP: single nucleotide polymorphism
LD: linkage disequilibrium

## Declarations

### Ethical Approval and Consent to participate

No human subjects were directly involved in this study. All the data used in this study was derived from existing de-identified biological samples from prior studies. Thus, ethical and patient consent was not required in this study.

## Acknowledgements

We would like to acknowledge the participants and investigators of the FinnGen project and the MEGASTROKE (International Stroke Genetics Consortium).

## Availability of supporting data

The data that support the findings of this study will be available from the corresponding author upon reasonable request.

## Authors’ contributions

M.Z. designed the study, developed the method, conducted data analysis, and wrote the manuscript. M.Z. supervised this project and is responsible for the overall content.

